# Genetic validation of the use of tocilizumab, statins and dexamethasone in COVID-19

**DOI:** 10.1101/2020.07.09.20149450

**Authors:** CM Schooling, SL Au Yeung, MK Kwok, JV Zhao

## Abstract

**Background:** New means of treating COVID-19 are urgently needed. Genetic validation of drugs can foreshadow trial results, and help prioritize investigations. We assessed whether common drugs, suggested as possible treatments for COVID-19 (tocilizumab, anakinra and statins) with established genetic proxies, are effective in COVID-19. We also included dexamethasone as a positive control exposure because the RECOVERY trial suggested benefit in severe COVID-19.

**Methods:** We assessed, using Mendelian randomization, whether genetic proxies of tocilizumab, anakinra, statins and dexamethasone use affected risk of very severe (cases=536, non-cases=329391) or hospitalized (cases=3199, non-cases=897488) COVID-19 using a recent genome-wide association study.

**Results:** Using rs2228145 (*IL6R*) to proxy effects of tocilizumab use, no association with very severe COVID-19 was found, but possibly an inverse association with hospitalized COVID-19 (odds ratio (OR) 0.83 per standardized effect of higher soluble interleukin-6r, 95% confidence interval 0.67 to 1.02). Using rs12916 (*HMGCR*) to proxy effects of statins use, an inverse association with very severe COVID-19 was found (OR 0.30 per standardized effect, 95% CI 0.10 to 0.89). Using rs6743376 and rs1542176 to proxy effects of anakinra use, no associations with COVID-19 were found. Dexamethasone, instrumented by cortisol, was possibly inversely associated with very severe COVID-19 (OR 0.20 per standardized effect 95% CI 0.04 to 1.04).

**Conclusion:** Our study provides some genetic validation for the use of both tocilizumab and statins in COVID-19, but not anakinra, whilst being consistent with the findings from the RECOVERY trial about dexamethasone. Investigation of the underlying mechanisms might facilitate re-purposing and development of effective treatments.

---

Many existing drugs have been suggested as possible candidates for treating COVID-19, but randomized controlled trials (RCTs) concerning their effects have not yet been completed. Evidence as to effectiveness in related conditions, such as acute respiratory distress syndrome (ARDS) or sepsis, is also limited. Moreover, the ‘cytokine storm’ that occurs in COVID-19 may differ from the clinical course of typical ARDS. The coagulopathy associated with COVID-19 also appears to differ from that typically seen in sepsis being more prothrombotic,^1 2^ with a higher risk of venous thromboembolism (VTE).^3^ In this situation where treatments are needed urgently, rapid repositioning of existing drugs could be helpful and can be facilitated by genetic validation.^4^ The recent availability of COVD-19 genome wide association studies (GWAS) meta-analysis (https://www.covid19hg.org/results/) means such genetic validation is now possible. Specifically, a preliminary assessment of likely drug efficacy can be made from the effect of a drug’s genetic proxy on COVID-19, using Mendelian randomization (MR), which has previously been used to foreshadow the results of several major RCTs.^4^

Here, we used MR to investigate the role of tocilizumab, anakinra and statins in COVID-19 because they have recently been suggested to be potentially useful for treating the severe ‘cytokine storm’ associated with COVID-19,^5^ and they also have well-established, published genetic proxies.^6-8^ Tocilizumab and anakinra are largely immune modulating drugs, targeting interleukin 6 (IL-6)or its receptor^6^ and interleukin 1 receptor antagonism (IL-1Ra),^7^ respectively. Genetic evidence also exists for a possible role of these two drugs in some cardiovascular diseases, with tocilizumab possibly protective^6^ and anakinra possibly harmful.^7^ In contrast, RCTs indicate that statins prevent and treat cardiovascular disease, as well as reducing all-cause mortality.^9-11^ Statins target cholesterol, but also have haemostatic and immunomodulatory properties,^12-14^ including possibly reducing thromboembolism^15 16^ which is increasingly seen as being relevant to COVID-19.^17^ Observationally, tocilizumab has shown some promise in severely ill COVID-19 patients,^18^ however results from RCTs, including the RECOVERY trial, are still pending. Less information is available for anakinra, but some promising small studies exist.^19 20^ Observationally statins reduce COVID-19 deaths.^21^ However, observational studies of drug use are open to bias by indication, and so are not a definitive guide.

Here, we applied genetic proxies for effects of using each of these three drugs (tocilizumab, anakinra and statins) to GWAS of COVID-19 to assess their effects on very serious COVID-19 and hospitalized COVID-19. For completeness, we also considered any COVID-19. Given dexamethasone has already shown success in preventing death in COVID-19 patients being ventilated or receiving oxygen (i.e., very serious COVID-19) in the RECOVERY trial <u>(NCT04381936</u>),^22^ we used dexamethasone, instrumented by its mechanistic target, cortisol,^23^ as a positive control exposure for very serious COVID-19. Given, dexamethasone may only be beneficial for COVID-19 patients with serious respiratory distress,^22^ we only expected dexamethasone to be effective in very serious COVID-19. COVID-19 is affected by major determinants of survival, i.e., smoking and body mass index (BMI),^24 25^ and mainly affects older people.^26^ As such, COVID-19 studies are open to selection bias as potentially they are studies of people who have survived both competing risk of COVID-19 and their genetic make-up.^27^ To address any such bias, we also used multivariable Mendelian randomization, adjusted for smoking and BMI, for drugs known to affect survival.^27^

## Methods

As previously, we used rs2228145 (*IL6R*) to proxy effects of tocilizumab use,^6^ in standardized effects of its target, i.e., raising soluble interleukin-6 receptor (sIL-6r).^28^ We used rs6743376 (*IL1F10*) and rs1542176 (*RNU6-1180P*) to proxy effects of anakinra use,^7^ in standardized effects of its target, i.e., raising IL-1Ra.^7^ We used rs12916 (*HMGCR*) to proxy effects of statin use,^8^ in standardized effects of its target, i.e., reducing low-density lipoprotein (LDL) cholesterol. We used one independent (r^2^<0.05) genetic variant, rs12589136, (*SERPINA6*) for morning plasma cortisol,^29^ in standardized effects of cortisol, because cortisol is the target of dexamethasone.^23^ We applied these variants to the most recent GWAS of COVID-19 (https://www.covid19hg.org/results/) (accessed 29^th^ June 2020) comparing very severe COVID-19 (cases=536, non-cases=329,391), hospitalized COVID-19 (n=3199, non-cases=897,488), and any COVID-19 (cases=6,696, non-cases=1,073,072) to the population. Case status for very severe COVID-19 was laboratory confirmed COVID-19 hospitalized with respiratory support or death. Case status for hospitalized COVID-19 was hospitalized with laboratory confirmed infection, hospitalization due to COVID-19-related symptoms or self-reported hospitalized COVID-19 positive. Case status for any COVID-19 was laboratory confirmed infection, doctor diagnosis or self-report. The GWAS was adjusted for age, sex, age^2^ and sex*age.

We obtained MR estimates for effects of these drugs on COVID-19 from Wald estimates (genetic variant on COVID-19 divided by genetic variant on drug target) with the standard error obtained from the first term of Fieller’s theorem.^30^ We meta-analyzed Wald estimates, as necessary, using inverse variance weighting. To adjust for smoking and BMI, we used published independent genetic instruments (r^2^<0.05) for smoking initiation^31^ and BMI.^32^ Genetic associations with ever smoking, BMI and the drug target were obtained from the UK Biobank summary statistics (http://www.nealelab.is/uk-biobank) adjusted for age, sex, age^2^, sex*age, sex*age^2^ and the first 20 principal components. We reported the multivariable conditional F-statistic as a measure of instrument strength and the multivariable Q-statistic as a measure of instrument pleiotropy,^33^ the latter potentially indicating genetic pleiotropy or selection bias. Given statins improve survival,^9-11^ to avoid selection bias from inevitably only selecting survivors of statins and other causes of COVID-19, we also adjusted statin estimates for smoking and BMI.^27^ No such evidence of effects on survival exists for tocilizumab, anakinra or dexamethasone so no such adjustment was made for them.

We used the MendelianRandomization R package to obtain univariable estimates, the MR-Base ld_clump R package to obtain independent genetic variants in the multivariable MR and the MVMR R package to obtain multivariable MR estimates, F-statistic and Q-statistic. This study of publicly available genetic summary statistics, obtained with consent, does not require ethical approval.

## Results

Genetically predicted effects of tocilizumab use had no effect on very severe COVID-19 but were in the direction of benefit for hospitalized COVID-19 and any COVID-19 (Figure 1). No genetically predicted effects of anakinra use were evident (Figure 1). Genetically predicted effects of statin use were protective for very severe COVID and with directionally similar estimates for hospitalized COVID, and for any COVID, when accounting for selection bias using multivariable MR (Figure 1 and Supplementary Table 1). The multivariable Q-statistic was not significant (Supplementary Table 1) and so did not indicate pleiotropy. As expected, genetically predicted effects of dexamethasone use for very severe COVID was in the direction of benefit, but not for hospitalized COVID or any COVID (Figure 1).

**Figure 1:**
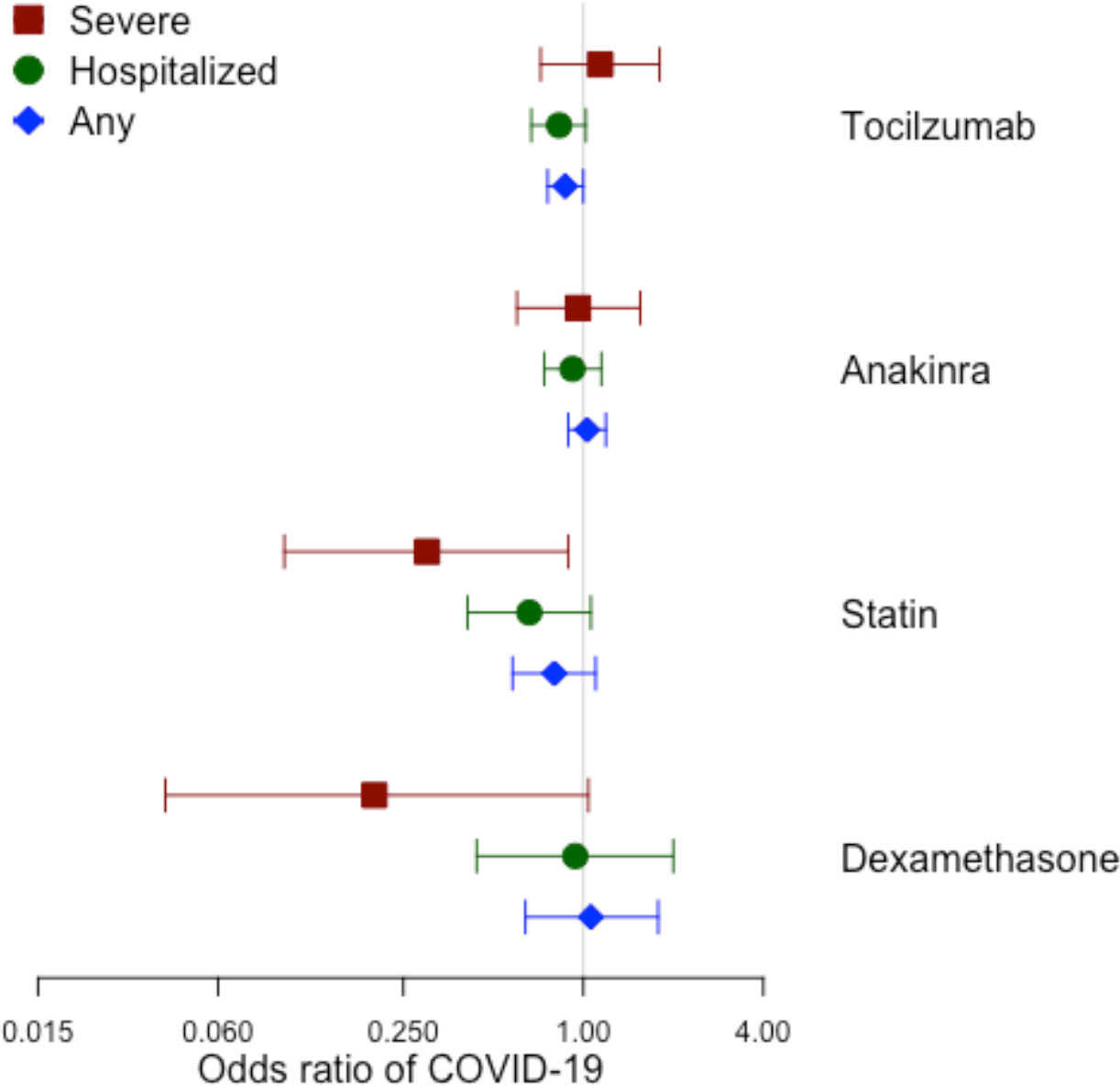
Mendelian randomization estimates (odds ratios with 95% confidence intervals) for effects of genetically predicted tocilizumab use,^6^ anakinra use,^7^ statin use,^8^ and dexamethasone use (proxied by cortisol)^29^ on severe, hospitalized and any COVID-19 from inverse variance weighting with fixed effects, with adjustment for smoking and BMI for statins.

## Discussion

Our study suggests that tocilizumab may be beneficial in hospitalized COVID-19 and possibly may prevent COVID-19. Our study also raises the possibility that statins may be valuable in treating very severe COVID-19. However, we found no indication that anakinra would be beneficial in COVID-19. Our study is also consistent with the RECOVERY trial in suggesting that dexamethasone may be protective in very severe COVID.^22^

Despite providing information that may be relevant to ameliorating the COVID-19 pandemic, this study has limitations. First, valid genetic instruments for MR should fulfil three assumptions, i.e., relate strongly to the effects of drug use, not be associated with potential confounders of the effects of drug use and satisfy the exclusion restriction assumption, i.e., only affect COVID-19 via the drug in question. The genetic variants used to predict effects of tocilizumab, anakinra and statins were originally selected as functionally relevant to the corresponding phenotype, i.e., sIL-6r, IL-1Ra and LDL-cholesterol, in genes that reflect each drug’s specific mechanism of action (*IL6R, IL1F10*, and *HMGCR*, respectively).^6-8 28^ As such, the genetic effect of the drug, encompassing the mechanism of action may extend beyond the target phenotype. Statins, for example, are well-known to have a wide range of effects beyond lipid modulation.^8^ Both effects via the target phenotype as well as “off-target” effects may contribute to a drug’s success in a new indication. MR studies can be confounded by population stratification. However, we used genetic associations from GWAS mainly comprising people of European ancestry (https://www.covid19hg.org/results/). We also rely on the integrity of the design of the underlying genetic studies to be free from bias, particularly that the non-cases come from the same underlying population as the cases, which is achieved by largely using existing cohort studies (https://www.covid19hg.org/results/). In contrast, a previous COVID-19 GWAS used blood donors as controls, who are likely to be younger and healthier than the general population which generated the COVID cases.^34^ Nevertheless, the underlying studies used here were recruited from the living, who may be selected survivors, so we cannot exclude the possibility of selection bias, hence our use of adjustment for statins,^27^ which affects survival.^9-11^ Canalization, i.e., buffering of genetic factors during development, may occur and obscure genetic effects, however COVID-19 is a new disease for which adaptions are unlikely to have occurred. Our findings, largely in Europeans, may not generalize to other populations. However, causes are unlikely to act differently in different populations, although the causal mechanisms may not be as relevant in all settings.^35^ We were not able to assess whether these drug effects on COVID-19 differed by sex, because sex-specific GWAS are not available, but could be important in the future when more COVID-19 cases have accumulated. However, given the sex bias in response to viral infections in general and specifically COVID-19,^36^ this could be an important consideration. Our study is clearly preliminary because of the small number of cases in the COVID-19 GWAS. However, the result for dexamethasone is broadly consistent with evidence from the RECOVERY trial, and this adds confidence to the study design, and provides valuable information for drugs with unclear evidence from RCTs.

Tocilizumab is an immunomodulatory agent indicated for treatment of rheumatoid arthritis (RA) and giant cell arteritis,^6^ with genetic evidence of possible benefit for aortic aneurysm.^6^ Anakinra is an effective treatment for RA, with genetic evidence of possible harm for ischemic heart disease (IHD),^7^ which might outweigh its beneficial immunomodulatory effects in COVID-19. Statins have pleiotropic effects, including on the immune and hemostatic systems.^12 13 37-39^ Statins and anakinra also have opposite effects on testosterone,^40^ when testosterone raises risk of VTE.^41^ Dexamethasone may also have pleiotropic effects on testosterone.^42^ Whether testosterone provides a unifying explanation for the effects of these different drugs on COVID-19 and sex-specific vulnerability to COVID-19^36^ remains to be determined. More pertinently, statins reduce thrombin generation,^43-46^ one of the few coagulation factors that increases risk of IHD.^47^ Given statins are an easily available, widely used and well-tolerated drug that might ameliorate COVID-19 via several mechanisms, trials of statins in COVID-19 might be valuable.

## Conclusion

Our study provides some genetic validation for the use of both tocilizumab and statins in COVID-19, but not for use of anakinra, whilst being consistent with the findings from the RECOVERY trial about dexamethasone. Further investigation of their role and underlying mechanisms could facilitate development of effective treatments for COVID-19.

## Data Availability

We only used publicly available data

## Acknowledgements

We are very grateful to *The COVID-19 host genetics initiative* for creating the COVID-19 GWAS and making summary statistics publicly available. We are also grateful to Ben Neale for making UK Biobank genetic summary statistics publicly available.

## Author Contributions

SLAY suggested the study concept. JVZ and CMS operationalized it with help from SLAY and MKK. CMS conducted the initial analysis, with help from JVZ. CMS wrote the first draft. All authors contributed to drafting and revising the article for intellectual content. All authors are accountable for all aspects of the work. All authors approved the final version.

## Competing Interest

This study had no funding. We have no support from any organization apart from governmental research funders. We have no other relationships that could have influenced this paper.

**Supplementary Table 1:**
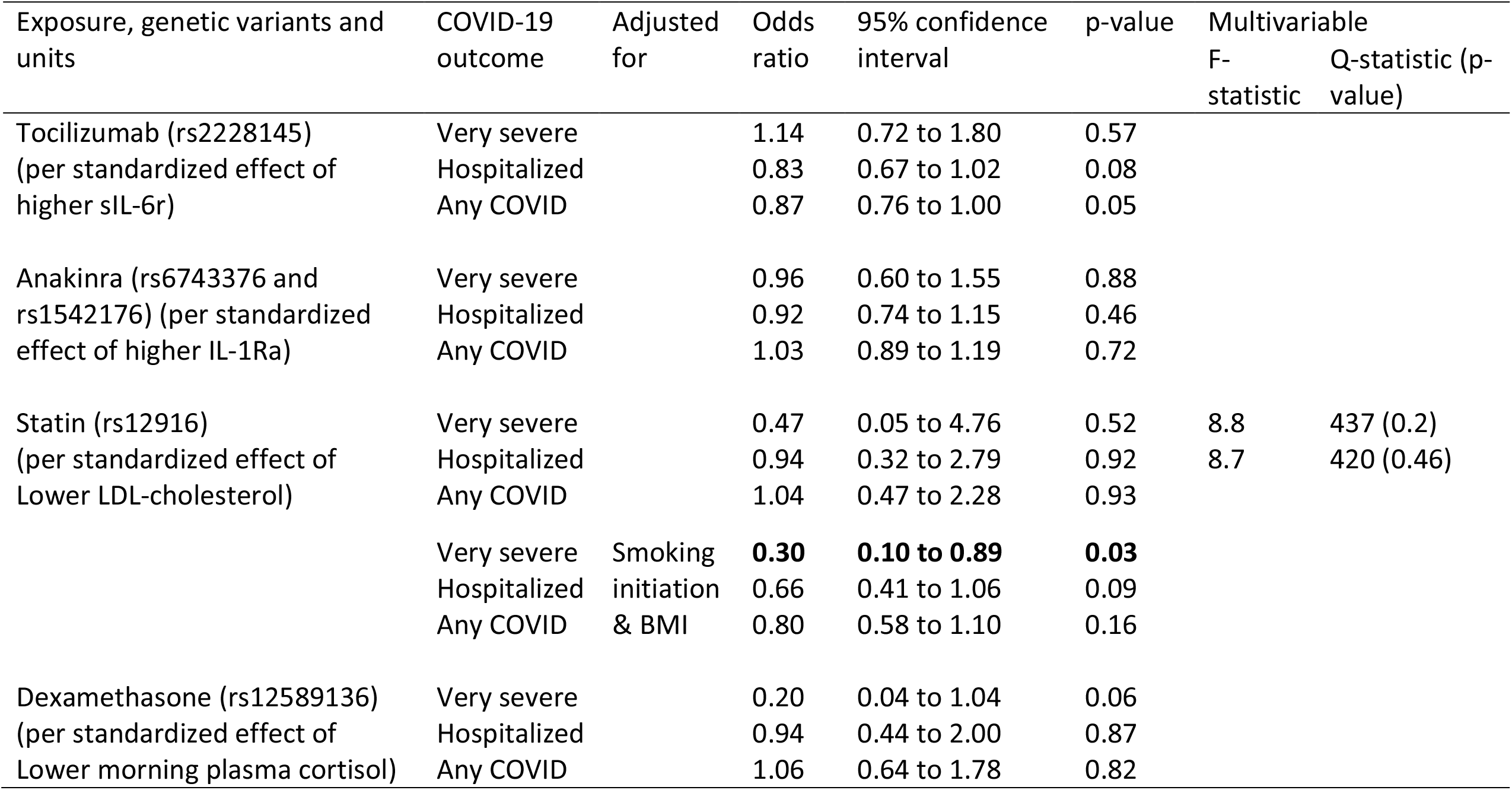
Mendelian randomization estimates for effects of genetically predicted tocilizumab use,^6^ anakinra use,^7^ statin use,^8^ and dexamethasone use (proxied by cortisol)^29^ on COVID-19 using inverse variance weighting with fixed effects.

## Notes

### Competing Interest Statement

The authors have declared no competing interest.

### Funding Statement

This work had no funding

### Author Declarations

This is a study of publicly available genetic summary data that does not require IRB approval.

